# Development of a COVID-19 Application Ontology for the ACT Network

**DOI:** 10.1101/2021.03.15.21253596

**Authors:** Shyam Visweswaran, Malarkodi J. Samayamuthu, Michele Morris, Griffin M. Weber, Douglas MacFadden, Philip Trevvett, Jeffrey G. Klann, Vivian Gainer, Barbara Benoit, Shawn N. Murphy, Lav Patel, Nebojsa Mirkovic, Yuliya Borovskiy, Robert D. Johnson, Matthew C. Wyatt, Amy Y. Wang, Robert W. Follett, Ngan Chau, Wenhong Zhu, Mark Abajian, Amy Chuang, Neil Bahroos, Phillip Reeder, Donglu Xie, Jennifer Cai, Elaina R. Sendro, Robert D. Toto, Gary S. Firestein, Lee M. Nadler, Steven E. Reis

## Abstract

Clinical data networks that leverage large volumes of data in electronic health records (EHRs) are significant resources for research on coronavirus disease 2019 (COVID-19). Data harmonization is a key challenge in seamless use of multisite EHRs for COVID-19 research. We developed a COVID-19 application ontology in the national Accrual to Clinical Trials (ACT) network that enables harmonization of data elements that that are critical to COVID-19 research. The ontology contains over 50,000 concepts in the domains of diagnosis, procedures, medications, and laboratory tests. In particular, it has computational phenotypes to characterize the course of illness and outcomes, derived terms, and harmonized value sets for SARS-CoV-2 laboratory tests. The ontology was deployed and validated on the ACT COVID-19 network that consists of nine academic health centers with data on 14.5M patients. This ontology, which is freely available to the entire research community on GitHub at https://github.com/shyamvis/ACT-COVID-Ontology, will be useful for harmonizing EHRs for COVID-19 research beyond the ACT network.

## 1 INTRODUCTION

On March 11, 2020, the coronavirus disease 2019 (COVID-19) caused by the severe acute respiratory syndrome coronavirus 2 (SARS-CoV-2) was declared a pandemic by the World Health Organization [1]. The striking transmissibility of SARS-CoV-2 has resulted in the rapid spread of COVID-19 worldwide that has severely strained healthcare delivery systems and public health agencies around the globe. Vast gaps in knowledge remain in understanding the natural course of the disease and characterization of risk factors of disease severity and poor outcomes. A significant source of data for COVID-19 research is patient data captured in electronic health records (EHRs). To leverage EHRs for COVID-19 research, several large clinical data networks including the Accrual to Clinical Trials (ACT) network [2], the National Patient-Centered Clinical Research Network (PCORnet) [3] and the All of Us Research Program [4] enhanced their capabilities to conduct COVID-19 EHR-based research. In addition, several dedicated consortia have been formed, including the National COVID-19 Cohort Collaborative (N3C) [5] and the Consortium for Clinical Characterization of COVID-19 by EHR (4CE) [6].

The ACT network is a federated network of Clinical and Translational Science Award (CTSA) sites that has implemented an efficient and extensible electronic infrastructure to transform clinical and translational research [2]. The goal of the ACT network is to enable investigators to perform cohort exploration, analyze data, and identify patients who are eligible for clinical trials on a national scale using EHR data. The network currently consists of EHR data on more than 150 million patients across 50 CTSA sites.

The technology underpinning the ACT network consists of local Informatics for Integrating Biology at the Bedside (i2b2) [7] and Observational Medical Outcomes Partnership (OMOP) [8] EHR repositories that are integrated by the Shared Health Research Information Network (SHRINE) platform [9]. The SHRINE platform facilitates the querying of EHRs in real time across the network with an easy-to-use query tool that is based on the i2b2 Query and Analysis Tool [10]. The SHRINE query tool employs a user-friendly query language that enables querying data across the sites in the network using medical terminologies where in each terminology the terms are arranged in a hierarchy for easy navigation. In the context of SHRINE, we call a terminology with hierarchical relations an application ontology (or simply ontology). The SHRINE ontologies have a tree-like hierarchical structure in which concepts closer to the root are more general than concepts located near the leaves. The tree-like structure enables the investigator to navigate the concepts and construct succinct queries by using the most general concepts that are applicable. The SHRINE query tool enables an investigator to drag and drop concepts from one or more ontologies to construct Boolean complex queries with operators such as negation, disjunction, and conjunction of concepts. Furthermore, temporal operators are available to define further constraints. The use of ontologies makes SHRINE a highly flexible network that facilitates addition of new concepts and entire ontologies based on research needs and new types of data.

Since the formation of the ACT network in 2015, the data harmonization work group has developed and distributed ontologies to the ACT sites. Each site installs the ACT ontologies and maps the concepts in the ACT ontologies to codes in the local i2b2 databases. Thus, the ACT ontologies achieve syntactic interoperability and local mappings achieve semantic interoperability across the network. The network is currently using ACT Ontology 2.0.1, which is based on the 2018AA release of the Unified Medical Language System (UMLS) Knowledge Sources [11]. The ontologies are in the domains of demographics, diagnoses (ICD-9-CM, ICD-10-CM), procedures (ICD-9-CM, ICD-10-PCS, CPT-4, HCPCS), medications (alphabetical, VA class), laboratory test results (LOINC), and visits. The ontologies are available for download at the ACT Network Wiki (https://dbmi-pitt.github.io/).

In response to the global pandemic caused by SARS-CoV-2, we mobilized the ACT network to support COVID-19 research at a national scale. To do so, we developed and deployed a COVID-19 application ontology and augmented EHR data to support the terms in the ontology. We believe that this ontology is accurate and useful not only to the ACT network but to other COVID-19 research efforts. However, we recognize that the ontology is likely incomplete and that there are limitations to the mapping of local codes and terms to ontology concepts that limit the comprehensiveness of the queries that can be constructed.

## 2 OBJECTIVE

Our objective was to develop and validate a COVID-19 ontology and to deploy it on the ACT network so that individual sites could augment the data to support the ontology. An important goal was to include concepts that would be particularly useful in COVID-19 research such as disease severity, events in the course of the disease, and outcomes.

## 3 METHODS

To obtain input from a diverse group of ACT members we communicated through weekly online meetings, a shared GitHub repository, and an i2b2 server dedicated for viewing the ontology as it was developed. We established the following process to develop, validate, and deploy the ontology:

1. Determining coverage: We identified concepts that would be needed to capture important aspects of COVID-19, based on which we iteratively determined broad areas of coverage.

2. Developing ontology: A draft ontology was developed that included emerging ICD-10, CPT-4, HCPCS, and LOINC codes, computable phenotypes to characterize the course of illness and outcomes, harmonized value sets for COVID-19 laboratory tests, and derived concepts. The draft ontology was loaded into the dedicated i2b2 server and iteratively modified using feedback.

3. Validity testing: We deployed the ontology on a COVID-19 ACT network that consisted of nine ACT sites. This allowed for testing the validity of the ontology and uncovering local codes and terms that were missing in the computable phenotypes and derived concepts as well as uncovering values in COVID-19 laboratory tests that needed harmonization.

4. Refining ontology: Using feedback from validity testing, we added additional concepts and codes to the computable phenotypes and derived concepts, and included additional values in the COVID-19 laboratory tests.

5. Implementation testing: After deployment of the refined ontology on the COVID-19 ACT network, we conducted ‘smoke’ tests and performed data characterization of key concepts in the ontology to ensure quality of the data at the sites.

We repeated the above process for the development and release of each new version of the ontology.

## 4 RESULTS

Since the beginning of the pandemic, we developed, released and deployed three versions of the ontology. The latest ontology, ACT COVID-19 Version 3, consists of 52,476 terms in the domains of diagnosis, procedures, medications, and laboratory tests. We included terms and their associated codes from commonly used terminologies that include ICD-10-CM, CPT-4, HCPCS, LOINC, and SNOMED-CT. In addition, we crafted computable phenotypes, derived concepts and harmonized value sets that are pertinent to COVID-19.

The COVID-19 ontology has several unique features to enable users to conveniently query with terms that are related to the course of illness and outcomes (see Figure 1). We highlight the following five categories of terms. 1) We identified and categorized emerging codes from ICD-10, CPT-4, HCPCS, and LOINC terminologies that were introduced in response to SARS-CoV-2. Table 1 lists the emerging codes that are included in the ontology. 2) We created **computable phenotypes** to characterize the course of illness and outcomes in COVID-19 that included illness severity [12], respiratory therapy management, and level of care. We developed computable phenotypes for three levels of illness severity – moderate, severe, and death – and for four levels of respiratory therapy management – supplemental oxygen, intubation, mechanical ventilation, and extracorporeal membrane oxygenation (ECMO) – and for each of these phenotypes we collected a set of relevant codes from ICD-10, CPT-4, and DRG. Example definitions of moderate illness and mechanical ventilation are shown in Table 2. Since there is variability across the sites in codes that are relevant to inferring illness severity, we iteratively identified and added codes based on the experience at the sites. 3) We created several **derived terms** and assigned them UMLS codes; for example, we included a “Moderate Illness (Derived)” concept with UMLS code C4740691 in the computable phenotype for moderate illness, and a “Mechanical Ventilation (Derived)” concept with UMLS code C0199470 in the computable phenotype for mechanical ventilation (see Table 2). These derived terms are useful in mapping data from EHRs of patients who are currently hospitalized and for which ICD-10 or CPT-4 codes may not be available since this coding typically occurs at or just after discharge. They are also useful for capturing additional local codes that are not part of the computable phenotype definition. 4) We created **harmonized value sets** for the growing number of SARS-CoV-2 nucleic acid antigen and antibody tests. The harmonized values are comprised of positive, negative, equivocal, and pending values and allow mapping of variously reported results to a set of four standardized values. The implementation of this mapping was a substantial undertaking at the participating sites and provided the key ability to query with standardized SARS-CoV-2 tests. 5) We identified terms in **existing** ACT ontologies that are likely to be useful for COVID-19 research and included them in the COVID-19 ontology for convenience. For example, we identified and added classes of medications that are relevant to COVID-19 research.

**Figure 1:**
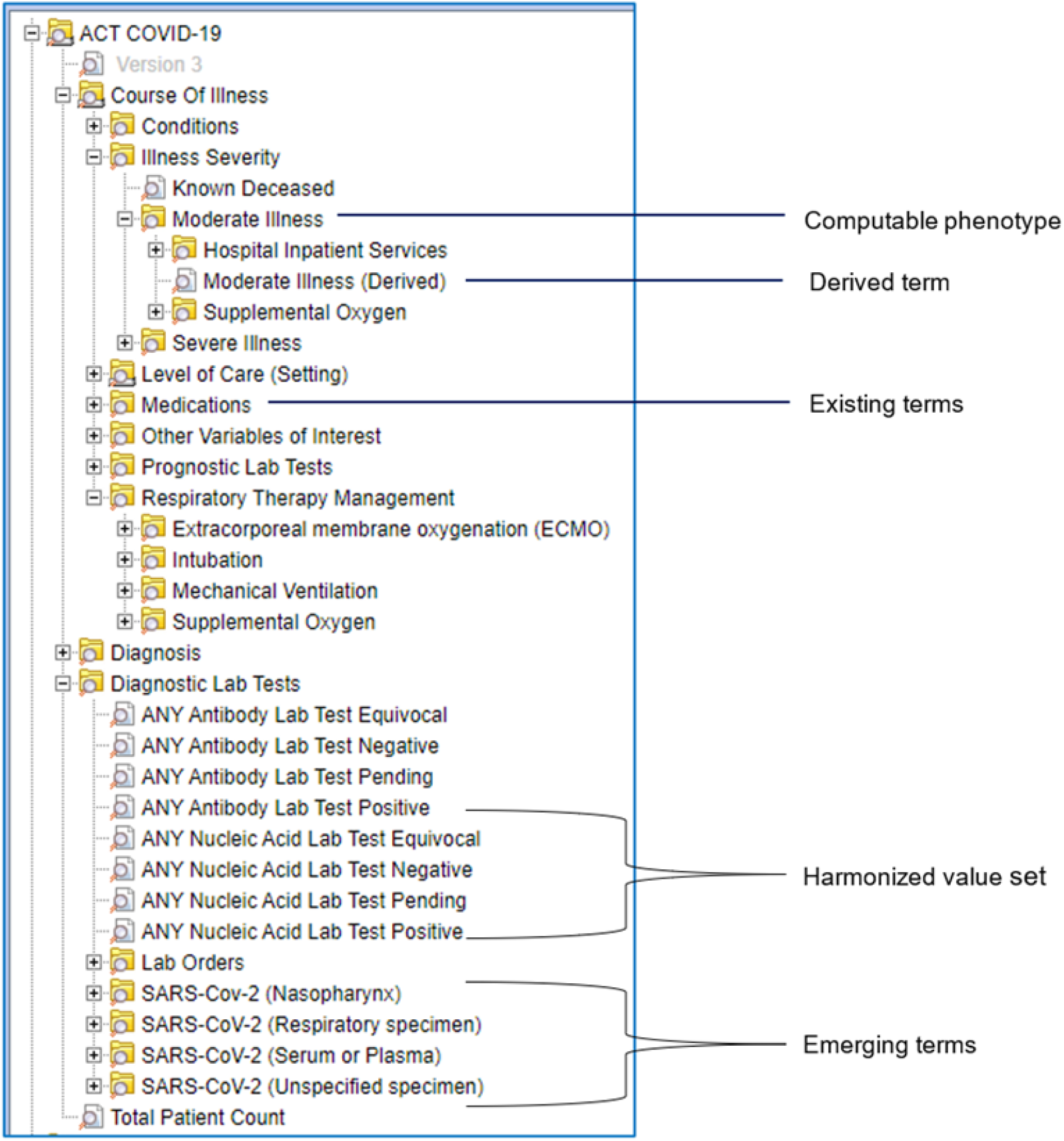
Screenshot of ACT COVID-19 ontology with illustrative examples of computable phenotype, derived term, existing terms, harmonized value set, and emerging terms. In the screenshot, only terms are displayed for easy comprehension; however, codes can also be displayed.

**Table 1:** Emerging SARS-CoV-2 codes in ICD-10, CPT-4, HCPCS, SNOMED-CT, and LOINC.

| Terminology | Codes |
| --- | --- |
| ICD-10 | U07.1, U07.2 |
| CPT-4 | 87635, 86318, 86328, 86769, 87426, 86408, 86409 |
| HCPCS | U0001, U0002 |
| SNOMED-CT | 3947185011, 120646007, 258603007, 430304001, 3947197012, 3947183016 |
| LOINC | 10 for laboratory test orders, 38 for SARS-CoV-2 nucleic acid antigen test results, 11 for SARS-CoV-2 antibody test results |

**Table 2:** Example computable phenotype definitions for moderate illness (a level of illness severity) and mechanical ventilation (a level of respiratory therapy management). Included in each of the computable phenotypes definitions is a derived term that is used for mapping data that has not been assigned standard codes.

| Computable phenotype | Codes |
| --- | --- |
| Moderate illness (illness severity) | Hospital Inpatient Services:<br>CPT-4: 1013659, 1013682, 1013683, 99238, 99239, 1013660, 1013661, 99223, 99222, 99221, 1013675, 99236, 99235, 99234, 1013668, 99233, 99231, 99232 |
|  | Supplemental Oxygen:<br>ICD-10-PCS: 3E0F7GC<br>UMLS: C4534306 |
|  | Moderate Illness (Derived term):<br>UMLS: C4740691 |
| Mechanical ventilation (respiratory therapy management) | Ventilation Diagnosis:<br>ICD-10-CM: J95.850, J95.851, J95.859, Z99.12<br>DRG: 475, 566 |
|  | Ventilation Procedures:<br>ICD-10-PCS: 5A09557, 5A09558, 5A09559, 5A0955A, 5A0955B, 5A0955Z<br>CPT-4: 94662, 94660, 94656, 94657, 94003, 94002 |
|  | Mechanical Ventilation (Derived term):<br>UMLS: C0199470 |

Counts of patients with SARS-CoV-2 nucleic acid laboratory test results from the ACT COVID-19 network are shown in Table 3. Preliminary feedback from investigators of the ACT COVID-19 network has been encouraging. Investigators found the computable phenotypes and the harmonized SARS-CoV-2 laboratory test values to be critical in identifying relevant cohorts.

**Table 3:** Counts of patients with SARS-CoV-2 nucleic acid and antibody laboratory test results from the ACT COVID-19 network.

| CTSA site | Total patient count | Nucleic acid laboratory test results |  |  | Antibody laboratory test results |  |  |  |  |
| --- | --- | --- | --- | --- | --- | --- | --- | --- | --- |
|  |  | Positive | Negative | Equivocal | Pending | Positive | Negative | Equivocal | Pending |
| Site 1 | 133,065 | 9,595 | 95,095 | 0 | 175 | 55 | 0 | 0 | 0 |
| Site 2 | 114,655 | 3,855 | 79,715 | 1,195 | 40 | 215 | 3,005 | 220 | 0 |
| Site 3 | 2,846,345 | 1,215 | 46,800 | 110 | 25 | 260 | 10670 | 90 | 45 |
| Site 4 | 3,578,620 | 11,010 | 87,080 | 405 | 530 | 0 | 0 | 0 | 0 |
| Site 5 | 2,962,235 | 8,465 | 90,490 | 0 | 0 | 185 | 2,275 | 0 | 0 |
| Site 6 | 889,640 | 235 | 840 | 0 | 0 | 0 | 0 | 0 | 0 |
| Site 7 | 3,234,770 | 3,025 | 46,440 | 0 | 0 | 175 | 1,755 | 0 | 0 |
| Site 8 | 717,180 | 4,030 | 48,720 | 35 | 0 | 0 | 0 | 0 | 0 |
| Site 9 | 85,345 | 2,710 | 66,535 | 225 | 15 | 900 | 13,130 | 0 | 0 |

## 5 DISCUSSION

In response to the pandemic, we mobilized the ACT network to develop and deploy a COVID-19 ontology to enable real-time cohort discovery across the ACT network. The development, implementation and validation steps described in the Methods section enabled us to rapidly develop and test a comprehensive COVID-19 ontology. We developed and released three versions of the ontology, evaluated the utility and the completeness of the computable phenotypes, assessed correctness of mapping of local codes and terms to the derived terms, and created a nine-site ACT COVID-19 network with access to the EHRs of 14.5M patients.

We have made the COVID-19 ontology files with accompanying documentation freely available to the entire research community on GitHub at https://github.com/shyamvis/ACT-COVID-Ontology. Furthermore, the ontology can be browsed freely at http://dbmi-ncats-test01.dbmi.pitt.edu/webclient/. We will continue to develop and release new versions at these websites.

A limitation of the ontology is that the inclusion of codes and terms in the computable phenotypes is based on a limited number of sites though we obtained feedback from nine large healthcare systems across the United States. Thus, the heterogeneity of coding that we observed is likely to be limited. As we deploy the ontology across all 50 sites in the ACT network, we will solicit feedback and update the ontology iteratively. Another limitation is that new codes for COVID-19 are being continually introduced, especially LOINC codes for new SARS-CoV-2 tests. Therefore, it is critical to keep the ontology updated with the newest codes. We plan to develop and publish new versions of the ontology on a regular basis.

Though we primarily developed the COVID-19 ontology for the ACT network, the ontology is has been leveraged by other COVID-19 research groups. In particular, the computable phenotypes that characterize the severity COVID-19 and disease outcomes have be beneficial in standardizing these computable phenotype definitions beyond the ACT network [12]. Since the publication of the ontology, the 4CE, the N3C, and the PCORnet consortia have adapted the ontology for their own use.

## 6 CONCLUSION

Multisite EHRs provide a unique source for rapid COVID-19 research using clinical data. We developed a COVID-19 ontology to enable harmonization and querying of EHRs. The ontology was validated on a nine-site network and is being deployed on the full ACT network. The ontology offers a foundational first step for harmonizing EHRs for research on COVID-19.

## Data Availability

The ontology files are freely available at GitHub.

https://github.com/shyamvis/ACT-COVID-Ontology

## ACKNOWLEDGEMENTS

This work was supported by the National Center for Advancing Translational Sciences of the National Institutes of Health under grant numbers UL1 TR000005-09S1 and UL1 TR001857-01S1, and by the National Library of Medicine of the National Institutes of Health under award numbers R01 LM012095. The content is solely the responsibility of the authors and does not necessarily represent the official views of the National Institutes of Health.

